# Knowledge and use of lactational amenorrhea as a family planning method among adolescent mothers in Uganda: a secondary analysis of Demographic and Health Surveys between 2006 and 2016

**DOI:** 10.1101/2021.06.17.21259067

**Authors:** Catherine Birabwa, Pamela Bakkabulindi, Solomon T Wafula, Peter Waiswa, Lenka Benova

**Affiliations:** School of Public Health, Makerere University College of Health Sciences, Kampala, Uganda; Health Support Initiatives, P.O Box 10364, Kampala, Uganda; Division of Global Health, Department of Public Health Sciences, Karolinska Institutet, Stockholm, Sweden; Institute of Tropical Medicine, Antwerp, Belgium

**Keywords:** Lactational Amenorrhea Method, adolescents, Uganda, contraceptive methods, breastfeeding, birth spacing

## Abstract

**Objective:** To assess the level of knowledge and use of lactational amenorrhea method (LAM) among adolescents in Uganda between 2006 and 2016 using nationally representative data from Demographic and Health Surveys (DHS).

**Design:** Cross-sectional design involving analysis of three DHS (2006, 2011, and 2016) in Uganda.

**Setting:** The data was collected in Uganda. The DHS are nationally representative surveys on a wide range of indicators including contraception knowledge and use.

**Participants:** A total of 8,250 adolescents (15-19 years) and 7,110 young women (20-24 years) were included.

**Primary outcome measure:** Use of LAM among adolescents and young women with a livebirth within six months before each survey.

**Results:** In 2016, less than 1% of eligible adolescents correctly used LAM and 56% were passively benefitting from LAM. The median duration of postpartum amenorrhea (PPA) among adolescents in 2016 was 6.9 months, declining from 8.3 months in 2006. Compared to adolescents, eligible young women had higher knowledge of LAM and higher medianPPA duration in 2016. The percentage of eligible adolescents who met the LAM criteria irrespective of whether they reported LAM use (protected by LAM) decreased from 76% in 2006 to 57% in 2016. More than 50% of eligible adolescents were aware of LAM in 2016, increasing from 6% in 2006, potentially in part due to change in survey question.

**Conclusion:** Despite increasing awareness of LAM, reported and correct use of LAM was low among adolescents who could benefit from this method in Uganda, and declining over time. Support for adolescents to harness the benefits of correct LAM use should be increased. Additional research is needed to better understand the dynamics of LAM use in adolescents, including the transition to use of other modern contraceptive methods.

**Strengths and limitations of this study:**

➢ Lactational amenorrhea method has the potential to promote healthy timing and spacing of pregnancies and to reduce repeat adolescent births, yet, its use among Ugandan adolescents has not been examined.
➢ This study provides useful insights into the behavior of adolescents mothers which can be targeted to improve their reproductive knowledge and wellbeing.
➢ The study used data from three standardised nationally representative surveys, thus findings are generalizable and comparable over time and across age groups.
➢ Assessment of knowledge of LAM was affected by a change in the phrasing of the question in 2016, thus providing limited comparability with previous surveys.
➢ The measurement of LAM use was based on self-report.

## INTRODUCTION

Reducing adolescent pregnancy rates is an important public health issue in sub-Saharan Africa (SSA), where high fertility and unmet need for family planning (FP) persist. Estimates show that 21 million adolescents aged 15-19 years in developing countries become pregnant every year (Neal et al., 2012, World Health Organization, 2020). Of these, 2.5 million are younger adolescents 12-15 years and at least 10 million are unintended pregnancies (Neal et al., 2012, World Health Organization, 2020). The prevalence of adolescent first births in SSA is estimated at 50% (Neal et al., 2012). As of 2019, about 218 million women in low- and middle-income countries (LMICs) had an unmet need for FP (Sully et al., 2020). In SSA, 25% of women aged 15-49 years are estimated to have an unmet need for FP (Ahinkorah et al., 2020). In 2019, 43% of adolescents in LMICs were estimated to have an unmet need for FP (Sully et al., 2020). Tackling the unmet need for FP among adolescents is critical especially in LMICs because it has significant implications on their reproductive health and wellbeing (Figaroa et al., 2020). Adolescent childbearing has been associated with multiple adverse health outcomes (Amongin et al., 2020a, Burke et al., 2018), and the literature also shows several challenges of the motherhood transition period for adolescents, which leave them disempowered with feelings of shame and embarrassment (Erfina et al., 2019, Mangeli et al., 2017, Watts et al., 2015). Specifically, concern has been raised about the unmet need for FP among postpartum women (Abraha et al., 2018). The acceptability and correct use of various FP methods among women during this period is affected by various socio-cultural and structural factors (Coomson and Manu, 2019, Joshi et al., 2020).

Lactational Amenorrhea Method (LAM) is a recognized modern contraceptive method where postpartum amenorrheic women depend on the contraceptive effect of breastfeeding within the first six months after childbirth (Van der Wijden and Manion, 2015). LAM provides 98% protection against pregnancy if 1) the woman’s menstrual period has not returned since childbirth, 2) the baby is fully or nearly fully breastfed, and 3) the baby is less than six months old (Van der Wijden and Manion, 2015, World Health Organization, 2018). The correct use of LAM provides an effective and affordable contraceptive option for breastfeeding women, while also providing an opportunity for linkage and transition of mothers to other FP methods and services. Furthermore, LAM use does not require replenishment of contraceptive supplies or a healthcare provider after appropriate LAM counselling is given. Therefore, LAM can play an important role in preventing unwanted pregnancies during the postpartum period and consequently maternal deaths (Alege et al., 2016). However, its effectiveness may be undermined by factors or practices that affect the three criteria, especially breastfeeding. Evidence shows low breastfeeding rates (Rahman et al., 2020, Yilmaz et al., 2016) and a higher likelihood of exclusive breastfeeding (EBF) discontinuation before infants reach 6 months among adolescent mothers (Jara-Palacios et al., 2015). A study of Nigerian adolescents found lower rates of early breastfeeding initiation and EBF compared to women 20-24 years (Benova et al., 2020). Education level, mode of delivery, antenatal care attendance, and postnatal breastfeeding counselling are some of the factors influencing breastfeeding practices among adolescents (Benova et al., 2020, Yilmaz et al., 2016).

Uganda has one of the highest total fertility rates (5.4 children per woman), maternal mortality rates of 336/100,000 livebirths (Uganda Bureau of Statistics and ICF, 2017), and the youngest population with 78% under 30 years (Ejang, 2020) in the world. Adolescent pregnancy rate is 25% with about 360,000 teenage pregnancies occurring annually (Ministry of Health, 2016, Uganda Bureau of Statistics and ICF, 2017). Evidence shows that over 40% of pregnancies among Ugandan adolescents are unintended (Nalwadda et al., 2019) and there is an increasing occurrence of repeat adolescent births (<20 years) estimated at 56%, with a prevalence of short-birth intervals of 5.4% (Amongin et al., 2020b). Unmet need for child spacing among women with adolescent births or repeat adolescent births has also been reported (Amongin et al., 2020a, Amongin et al., 2020b). Lack of knowledge and access to general health services and youth-friendly FP care, in particular, are some of the key challenges affecting adolescent health in Uganda (Atuyambe et al., 2015). Evidence on use of LAM in adolescents in LMICs is still scarce. There is a need to better understand the duration, knowledge, and correct use of LAM as a postpartum FP method among adolescents (Figaroa et al., 2020).

The main objective of this study was to assess the knowledge and use of LAM among adolescents in Uganda between 2006 and 2016 using nationally representative data from Demographic and Health Surveys (DHS).

## METHODOLOGY

### Data

This study is a secondary analysis of the cross-sectional data collected during Uganda DHS in 2006, 2011, and 2016. These are nationally representative household surveys carried out every five years. The DHS employs a stratified two-stage cluster sampling design to select participating households. All women of reproductive age (15-49 years) who are either permanent residents of the selected households or visitors who stayed in the household the night before the survey are eligible to be interviewed. The women interviewed provide, among others, self-reported information on their education, marital status, reproductive history, family planning use, maternal health-seeking for recent live births, and breastfeeding practices. Data were collected by trained data collectors using pre-tested tools.

### Study population

Our analysis sample included women of reproductive age (15-49 years) at the time of each survey. The primary study population was adolescent girls aged 15-19 years, who were compared to young women aged 20-24 years. Within this sample, we also included a sub-group of adolescent girls and young women with a live birth in the six months before each survey as they are the population benefitting from LAM.

### Definitions

We estimated the following indicators capturing three dimensions of lactational amenorrhea: 1) Knowledge of LAM; 2) Use of LAM – including reported, correct, passive, and overall protection by lactational amenorrhea; and 3) Duration of postpartum amenorrhea (Table 1).

**Table 1.** Definitions of key study variables.

| Indicator (estimate) | Definition (numerator/ denominator) | Stratification by woman's age group (at time of survey) |
| --- | --- | --- |
| 1 Knowledge of LAM as FP method among all women (% with 95%CI) | Women who report ever hearing of LAM / All women | 15-19.9<br>20-24.9<br>All women 15-49 |
| 2 Knowledge of LAM among eligible women (% with 95%CI) | Women who report ever hearing of LAM / Women with a child <6 months of age | 15-19.9<br>20-24.9<br>All women 15-49 |
| 3 Current reported use of LAM as FP method among all women (% with 95%CI) | Women who report using LAM as their FP method at time of survey / All women | 15-19.9<br>20-24.9<br>All women 15-49 |
| 4 Current reported use of LAM as FP method among eligible women (% with 95%CI) | Women who report using LAM as their FP method at time of survey / Women with a child <6 months of age | 15-19.9<br>20-24.9<br>All women 15-49 |
| 5 Correct use of LAM among eligible women (% with 95%CI) | Women who report using LAM and meet the three criteria/ Women with a child <6 months of age | 15-19.9<br>20-24.9<br>All women 15-49 |
| 6 Passive use of LAM among eligible women (% with 95%CI) | Women who meet the three LAM criteria but do not report using LAM / Women with a child <6 months of age | 15-19.9<br>20-24.9<br>All women 15-49 |
| 7 Protection by LAM among all women (% with 95%CI) | Women who meet the three LAM criteria regardless of whether they report the use of LAM or not / All women | 15-19.9<br>20-24.9<br>All women 15-49 |
| 8 Protection by LAM among eligible women (% with 95%CI) | Women who meet the three LAM criteria regardless of whether they report the use of LAM or not / Women with a child <6 months of age | 15-19.9<br>20-24.9<br>All women 15-49 |
| 9 Duration of postpartum amenorrhea in months (median +IQR) | Number of months before menstrual period returned after most recent birth in the 36 months before the survey | 15-19.9<br>20-24.9<br>All women 15-49 |

LAM=Lactational Amenorrhea Method, FP=family planning

***Knowledge of LAM*** was defined as the respondent being aware of LAM as a method of family planning, and was measured based on respondents’ binary (yes, no) answer to “*Have you ever heard of lactational amenorrhea as a family planning method?*” It should be noted that, in the 2016 survey, this question was followed with a probe that describes LAM further, that is, “*Up to six months after childbirth, before the menstrual period has returned, women use a method requiring frequent breastfeeding day and night*”. This probe was not used in the 2006 and 2011 surveys. We assessed knowledge of LAM among all women and among eligible women who could have been using this method at the time of the survey (those with infants below six months of age at the time of each survey).

***Use of LAM*** was based on the reported current use of lactational amenorrhea as a family planning method at the time of each survey (yes or no). This was measured by asking women whether they were doing anything or using any method to delay or avoid getting pregnant; and if so, what method they were using (multiple responses were allowed, for example, LAM and male condoms). We examined the use of LAM through four indicators:

1. Current use of LAM - women who reported using LAM as their method of family planning at the time of the survey. We applied two denominators: all women and eligible women (those with infants below six months at the time of the survey).
2. Correct use of LAM – women who reported LAM use and met the three LAM criteria, among eligible women (those with infants below six months at the time of the survey). The three criteria for correct use of LAM were; i) woman has a child <6months of age at the time of the survey, ii) woman’s menstrual period had not returned since the most recent birth, and iii) her infant was being exclusively breastfed at the time of the survey. *Exclusive breastfeeding* was defined as infants <6 months of age at the time of the survey who were fed exclusively with breastmilk, that is not given anything other than breastmilk in the 24 hours before the survey, except for oral medications.
3. Passive use of LAM - women meeting the three LAM criteria but who did not report LAM use, among eligible women (those with infants <6 months at the time of the survey).
4. Protection by LAM – women who met the three LAM criteria, regardless of whether they reported LAM use or not), among all women and eligible women.

#### Duration of postpartum amenorrhea

Postpartum amenorrhea (PPA) period was defined as the time to the resumption of menstruation period following childbirth. This was captured by calculating the median duration in months using the child recode file, following the standard DHS method (Tesfayi et al., 2008).

#### Other variables

We used data on women’s socio-demographic characteristics at the time of the surveys, including women’s age in years, marital status, education status, place and region of residence, and household wealth quintile. Marital status was categorized as “never in union”, currently in union/living with a man, and formerly in union/living with a man. Education was categorized as no education, primary education, secondary education, and higher education. Participants’ place of residence was either urban or rural, and household wealth quintiles as produced by the DHS.

### Statistical Analysis

Analysis was conducted using STATA 14 (StataCorp, Texas, USA). We utilized the *svyset* command in STATA to adjust for the sampling design used and for non-response. Descriptive statistics including frequencies and percentages were used to summarize categorical data while continuous data (duration of PPA) was expressed in terms of median and inter-quartile range. We computed the unweighted numbers of women for the given variable and the weighted percentages and 95% confidence interval. For all denominators using eligible women (with infants below six months at the time of the survey), we only included women with infants who were alive at the time of the survey due to breastfeeding as a criterion for correct LAM use. In the computation of reported use of LAM at the time of the survey, we assumed that all women who did not report using LAM were indeed not using LAM (missing values were recorded as 0). There was no missing data on other key variables for the study samples included in the analysis.

### Ethical approval

Data used for the analysis of this paper were obtained with permission from the Demographic and Health Survey Program (DHS) program of the US Agency for International Development. The DHS receives government permission, uses informed consent, and assures respondents of confidentiality. We did not require an ethics approval for this study since it involved secondary data analysis.

## RESULTS

### Socio-demographic characteristics of the study samples

The total number of adolescents included in the study was 8,250 and 7,110 young women (Table 2). Across the decade, primary education was the highest level attained in both adolescents (at an average 65%) and young women (∼ 55%) (Table 3). Approximately 77% of adolescents had never been married as compared to approximately 20% in the young women. Table 3 also shows an increasing trend in the proportion of adolescents (17.7% to 24.3%) or young women (22.0% to 29.9%) living in urban areas.

**Table 2.**
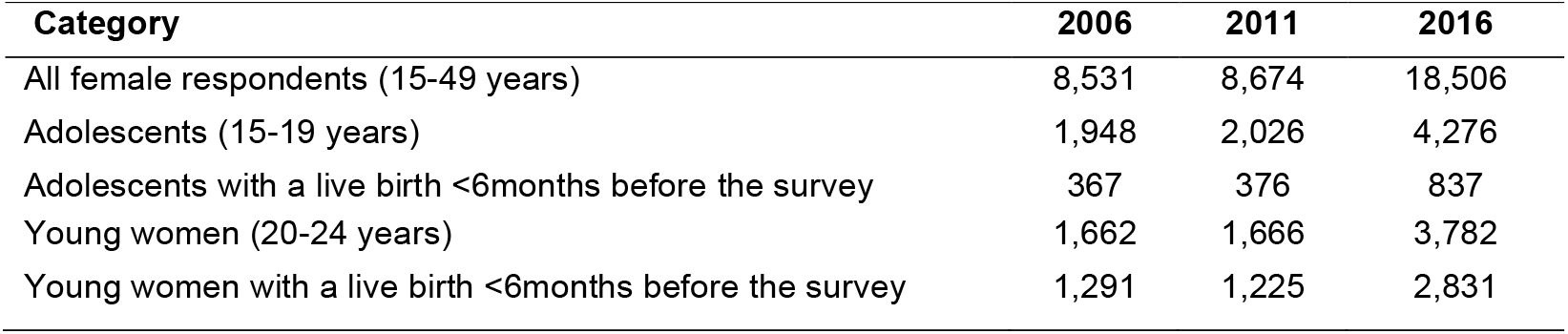
Sample size of women according to age group at the time of each survey and recent birth history, by survey.

**Table 3:**
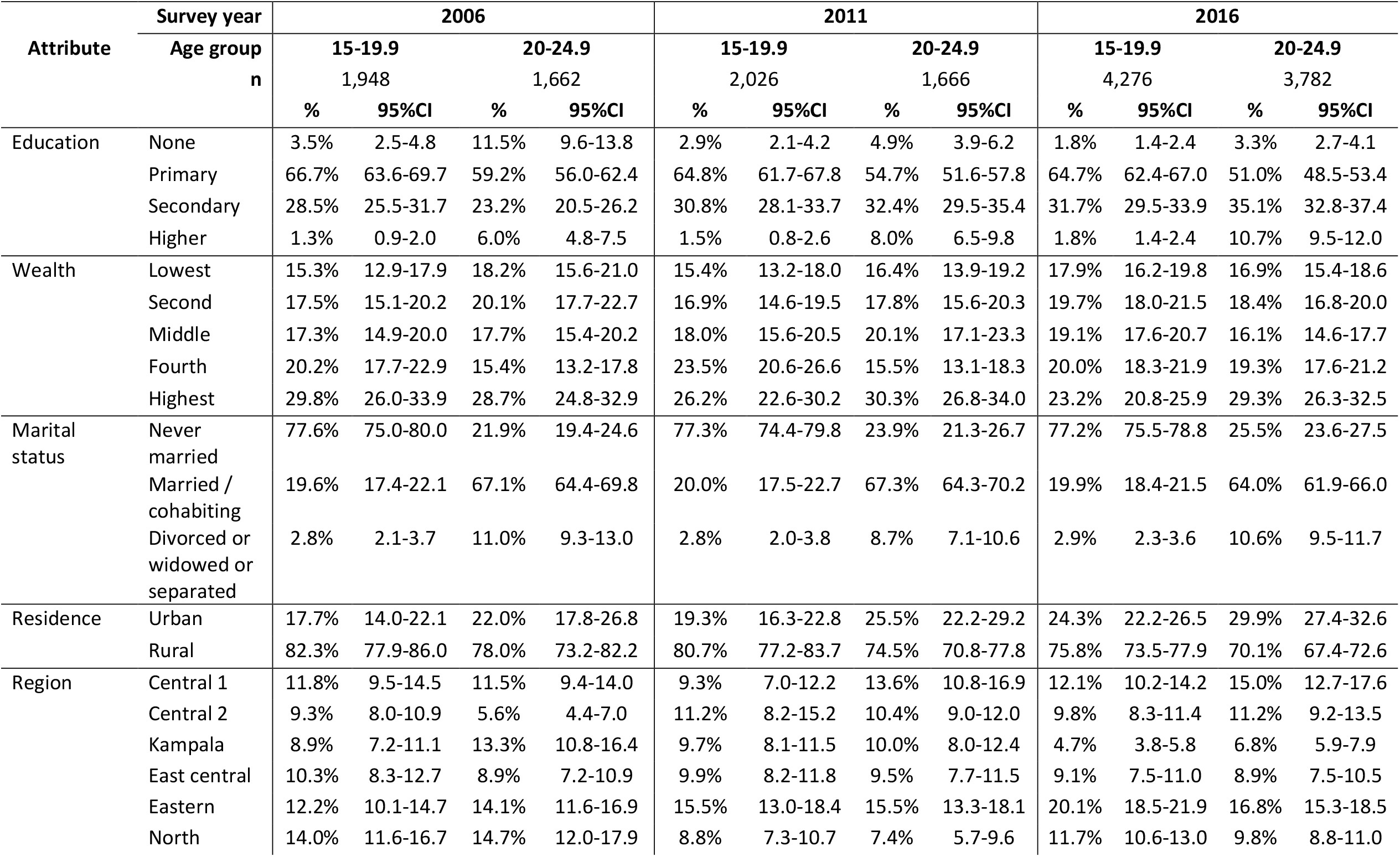
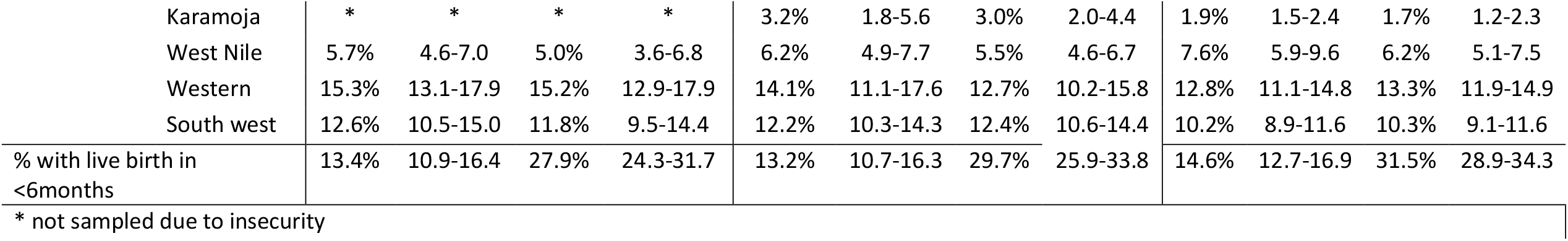
Socio-demographic characteristics of the study samples for UDHS 2006, 2011 and 2016.

| Attribute | Survey year | 2006 |  |  |  | 2011 |  |  |  | 2016 |  |  |  |
| --- | --- | --- | --- | --- | --- | --- | --- | --- | --- | --- | --- | --- | --- |
|  | Age group | 15-19.9 |  | 20-24.9 |  | 15-19.9 |  | 20-24.9 |  | 15-19.9 |  | 20-24.9 |  |
|  | n | 1,948 |  | 1,662 |  | 2,026 |  | 1,666 |  | 4,276 |  | 3,782 |  |
|  |  | % | 95%CI | % | 95%CI | % | 95%CI | % | 95%CI | % | 95%CI | % | 95%CI |
| Education | None | 3.5% | 2.5-4.8 | 11.5% | 9.6-13.8 | 2.9% | 2.1-4.2 | 4.9% | 3.9-6.2 | 1.8% | 1.4-2.4 | 3.3% | 2.7-4.1 |
|  | Primary | 66.7% | 63.6-69.7 | 59.2% | 56.0-62.4 | 64.8% | 61.7-67.8 | 54.7% | 51.6-57.8 | 64.7% | 62.4-67.0 | 51.0% | 48.5-53.4 |
|  | Secondary | 28.5% | 25.5-31.7 | 23.2% | 20.5-26.2 | 30.8% | 28.1-33.7 | 32.4% | 29.5-35.4 | 31.7% | 29.5-33.9 | 35.1% | 32.8-37.4 |
|  | Higher | 1.3% | 0.9-2.0 | 6.0% | 4.8-7.5 | 1.5% | 0.8-2.6 | 8.0% | 6.5-9.8 | 1.8% | 1.4-2.4 | 10.7% | 9.5-12.0 |
| Wealth | Lowest | 15.3% | 12.9-17.9 | 18.2% | 15.6-21.0 | 15.4% | 13.2-18.0 | 16.4% | 13.9-19.2 | 17.9% | 16.2-19.8 | 16.9% | 15.4-18.6 |
|  | Second | 17.5% | 15.1-20.2 | 20.1% | 17.7-22.7 | 16.9% | 14.6-19.5 | 17.8% | 15.6-20.3 | 19.7% | 18.0-21.5 | 18.4% | 16.8-20.0 |
|  | Middle | 17.3% | 14.9-20.0 | 17.7% | 15.4-20.2 | 18.0% | 15.6-20.5 | 20.1% | 17.1-23.3 | 19.1% | 17.6-20.7 | 16.1% | 14.6-17.7 |
|  | Fourth | 20.2% | 17.7-22.9 | 15.4% | 13.2-17.8 | 23.5% | 20.6-26.6 | 15.5% | 13.1-18.3 | 20.0% | 18.3-21.9 | 19.3% | 17.6-21.2 |
|  | Highest | 29.8% | 26.0-33.9 | 28.7% | 24.8-32.9 | 26.2% | 22.6-30.2 | 30.3% | 26.8-34.0 | 23.2% | 20.8-25.9 | 29.3% | 26.3-32.5 |
| Marital status | Never married | 77.6% | 75.0-80.0 | 21.9% | 19.4-24.6 | 77.3% | 74.4-79.8 | 23.9% | 21.3-26.7 | 77.2% | 75.5-78.8 | 25.5% | 23.6-27.5 |
|  | Married / cohabiting | 19.6% | 17.4-22.1 | 67.1% | 64.4-69.8 | 20.0% | 17.5-22.7 | 67.3% | 64.3-70.2 | 19.9% | 18.4-21.5 | 64.0% | 61.9-66.0 |
|  | Divorced or widowed or separated | 2.8% | 2.1-3.7 | 11.0% | 9.3-13.0 | 2.8% | 2.0-3.8 | 8.7% | 7.1-10.6 | 2.9% | 2.3-3.6 | 10.6% | 9.5-11.7 |
| Residence | Urban | 17.7% | 14.0-22.1 | 22.0% | 17.8-26.8 | 19.3% | 16.3-22.8 | 25.5% | 22.2-29.2 | 24.3% | 22.2-26.5 | 29.9% | 27.4-32.6 |
|  | Rural | 82.3% | 77.9-86.0 | 78.0% | 73.2-82.2 | 80.7% | 77.2-83.7 | 74.5% | 70.8-77.8 | 75.8% | 73.5-77.9 | 70.1% | 67.4-72.6 |
| Region | Central 1 | 11.8% | 9.5-14.5 | 11.5% | 9.4-14.0 | 9.3% | 7.0-12.2 | 13.6% | 10.8-16.9 | 12.1% | 10.2-14.2 | 15.0% | 12.7-17.6 |
|  | Central 2 | 9.3% | 8.0-10.9 | 5.6% | 4.4-7.0 | 11.2% | 8.2-15.2 | 10.4% | 9.0-12.0 | 9.8% | 8.3-11.4 | 11.2% | 9.2-13.5 |
|  | Kampala | 8.9% | 7.2-11.1 | 13.3% | 10.8-16.4 | 9.7% | 8.1-11.5 | 10.0% | 8.0-12.4 | 4.7% | 3.8-5.8 | 6.8% | 5.9-7.9 |
|  | East central | 10.3% | 8.3-12.7 | 8.9% | 7.2-10.9 | 9.9% | 8.2-11.8 | 9.5% | 7.7-11.5 | 9.1% | 7.5-11.0 | 8.9% | 7.5-10.5 |
|  | Eastern | 12.2% | 10.1-14.7 | 14.1% | 11.6-16.9 | 15.5% | 13.0-18.4 | 15.5% | 13.3-18.1 | 20.1% | 18.5-21.9 | 16.8% | 15.3-18.5 |
|  | North | 14.0% | 11.6-16.7 | 14.7% | 12.0-17.9 | 8.8% | 7.3-10.7 | 7.4% | 5.7-9.6 | 11.7% | 10.6-13.0 | 9.8% | 8.8-11.0 |

|  |  |  |  |  |  |  |  |  |  |  |  |  |
| --- | --- | --- | --- | --- | --- | --- | --- | --- | --- | --- | --- | --- |
| Karamoja | * | * | * | * | 3.2% | 1.8-5.6 | 3.0% | 2.0-4.4 | 1.9% | 1.5-2.4 | 1.7% | 1.2-2.3 |
| West Nile | 5.7% | 4.6-7.0 | 5.0% | 3.6-6.8 | 6.2% | 4.9-7.7 | 5.5% | 4.6-6.7 | 7.6% | 5.9-9.6 | 6.2% | 5.1-7.5 |
| Western | 15.3% | 13.1-17.9 | 15.2% | 12.9-17.9 | 14.1% | 11.1-17.6 | 12.7% | 10.2-15.8 | 12.8% | 11.1-14.8 | 13.3% | 11.9-14.9 |
| South west | 12.6% | 10.5-15.0 | 11.8% | 9.5-14.4 | 12.2% | 10.3-14.3 | 12.4% | 10.6-14.4 | 10.2% | 8.9-11.6 | 10.3% | 9.1-11.6 |
| % with live birth in<br><6months | 13.4% | 10.9-16.4 | 27.9% | 24.3-31.7 | 13.2% | 10.7-16.3 | 29.7% | 25.9-33.8 | 14.6% | 12.7-16.9 | 31.5% | 28.9-34.3 |

\* not sampled due to insecurity

### Knowledge of LAM

Knowledge of LAM as a FP method was analyzed for two categories of women; all women and eligible women (who had a child <6 months at the time of the survey). Between 2006 and 2016, knowledge of LAM among all and eligible adolescent girls increased. Among all adolescents, knowledge of LAM increased from 3.7% in 2006 to 36.9% in 2016; whilst among the eligible adolescents, it increased from 6.0% to 56.7%, respectively (Table 4). Compared to young women, knowledge of LAM among all or eligible women was generally lower among the adolescents. Among eligible young women, knowledge of LAM increased from 9.7% in 2006 to 64.1% in 2016.

### Use of LAM

We determined the use of LAM using four broad indicators: current reported use, correct use, passive use of LAM, and protection by LAM. The overall use of LAM over the ten years was low among adolescents, young women and all women of reproductive age. The overall current reported use or correct use of LAM was less than 2% (Table 4). The percentage of eligible adolescents or young women correctly using LAM was lower than the percentage who reported using LAM as their FP method. Among eligible women, the current reported use as of LAM in adolescents was 1.3% in 2016 as compared to 0.7% in young women; whilst the correct use of LAM in 2016 was 0.9% in adolescents compared to 0.5% in young women. Table 4 further shows that across the three surveys, the percentage of eligible women passively using LAM was higher compared to those reporting intentional LAM use (>50% of eligible women in all surveys and age groups). Also, the passive use of LAM decreased among all age groups over the review period. Among adolescent girls, passive LAM use decreased from 76.4% in 2006 to 56.3% in 2016, compared to young women among whom it decreased from 84.9% to 56.9% respectively. Between 2006 and 2016, the percentage of eligible women protected by LAM decreased across all age groups. As of 2016, at least 50% of women eligible for LAM were indeed protected by LAM in all three age groups, more so among all women of reproductive age at 62.0%. The percentage of eligible adolescents protected by LAM decreased from 76.4% in adolescents in 2006 to 57.2% in 2016, as compared to young women where it decreased from 84.9% in 2006 to 57.3% in 2016 (table 4).

**Table 1.**
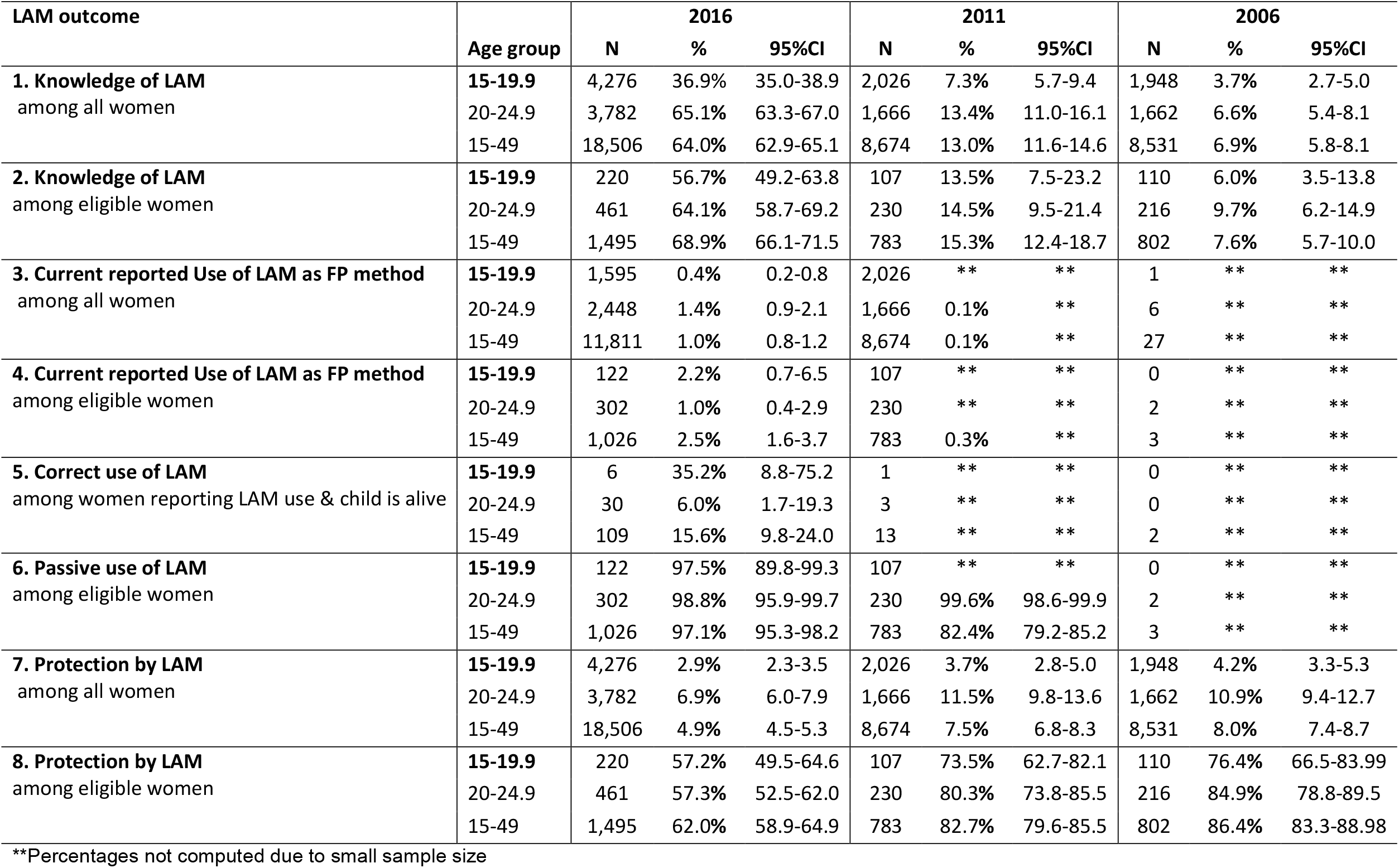
Knowledge and use of lactational amenorrhea and duration of postpartum amenorrhea, by age group and survey.

| LAM outcome | Age group | 2016 |  |  | 2011 |  |  | 2006 |  |  |
| --- | --- | --- | --- | --- | --- | --- | --- | --- | --- | --- |
|  |  | N | % | 95%CI | N | % | 95%CI | N | % | 95%CI |
| **1. Knowledge of LAM**<br>among all women | **15-19.9** | 4,276 | 36.9% | 35.0-38.9 | 2,026 | 7.3% | 5.7-9.4 | 1,948 | 3.7% | 2.7-5.0 |
|  | 20-24.9 | 3,782 | 65.1% | 63.3-67.0 | 1,666 | 13.4% | 11.0-16.1 | 1,662 | 6.6% | 5.4-8.1 |
|  | 15-49 | 18,506 | 64.0% | 62.9-65.1 | 8,674 | 13.0% | 11.6-14.6 | 8,531 | 6.9% | 5.8-8.1 |
| **2. Knowledge of LAM**<br>among eligible women | **15-19.9** | 220 | 56.7% | 49.2-63.8 | 107 | 13.5% | 7.5-23.2 | 110 | 6.0% | 3.5-13.8 |
|  | 20-24.9 | 461 | 64.1% | 58.7-69.2 | 230 | 14.5% | 9.5-21.4 | 216 | 9.7% | 6.2-14.9 |
|  | 15-49 | 1,495 | 68.9% | 66.1-71.5 | 783 | 15.3% | 12.4-18.7 | 802 | 7.6% | 5.7-10.0 |
| **3. Current reported Use of LAM as FP method**<br>among all women | **15-19.9** | 1,595 | 0.4% | 0.2-0.8 | 2,026 | ** | ** | 1 | ** | ** |
|  | 20-24.9 | 2,448 | 1.4% | 0.9-2.1 | 1,666 | 0.1% | ** | 6 | ** | ** |
|  | 15-49 | 11,811 | 1.0% | 0.8-1.2 | 8,674 | 0.1% | ** | 27 | ** | ** |
| **4. Current reported Use of LAM as FP method**<br>among eligible women | **15-19.9** | 122 | 2.2% | 0.7-6.5 | 107 | ** | ** | 0 | ** | ** |
|  | 20-24.9 | 302 | 1.0% | 0.4-2.9 | 230 | ** | ** | 2 | ** | ** |
|  | 15-49 | 1,026 | 2.5% | 1.6-3.7 | 783 | 0.3% | ** | 3 | ** | ** |
| **5. Correct use of LAM**<br>among women reporting LAM use & child is alive | **15-19.9** | 6 | 35.2% | 8.8-75.2 | 1 | ** | ** | 0 | ** | ** |
|  | 20-24.9 | 30 | 6.0% | 1.7-19.3 | 3 | ** | ** | 0 | ** | ** |
|  | 15-49 | 109 | 15.6% | 9.8-24.0 | 13 | ** | ** | 2 | ** | ** |
| **6. Passive use of LAM**<br>among eligible women | **15-19.9** | 122 | 97.5% | 89.8-99.3 | 107 | ** | ** | 0 | ** | ** |
|  | 20-24.9 | 302 | 98.8% | 95.9-99.7 | 230 | 99.6% | 98.6-99.9 | 2 | ** | ** |
|  | 15-49 | 1,026 | 97.1% | 95.3-98.2 | 783 | 82.4% | 79.2-85.2 | 3 | ** | ** |
| **7. Protection by LAM**<br>among all women | **15-19.9** | 4,276 | 2.9% | 2.3-3.5 | 2,026 | 3.7% | 2.8-5.0 | 1,948 | 4.2% | 3.3-5.3 |
|  | 20-24.9 | 3,782 | 6.9% | 6.0-7.9 | 1,666 | 11.5% | 9.8-13.6 | 1,662 | 10.9% | 9.4-12.7 |
|  | 15-49 | 18,506 | 4.9% | 4.5-5.3 | 8,674 | 7.5% | 6.8-8.3 | 8,531 | 8.0% | 7.4-8.7 |
| **8. Protection by LAM**<br>among eligible women | **15-19.9** | 220 | 57.2% | 49.5-64.6 | 107 | 73.5% | 62.7-82.1 | 110 | 76.4% | 66.5-83.99 |
|  | 20-24.9 | 461 | 57.3% | 52.5-62.0 | 230 | 80.3% | 73.8-85.5 | 216 | 84.9% | 78.8-89.5 |
|  | 15-49 | 1,495 | 62.0% | 58.9-64.9 | 783 | 82.7% | 79.6-85.5 | 802 | 86.4% | 83.3-88.98 |

\*\*Percentages not computed due to small sample size

### Duration of postpartum amenorrhea

Over the ten-year period, the median duration of postpartum amenorrhea declined among both adolescents and young women who had had a live birth in the 3 years preceding each survey (Figure 1). Among the adolescents, the median duration of PPA decreased from 8.3 months in 2006 to 6.9 months in 2016, while in young women it decreased from 11.2 months to 8.0 months respectively. Across all age groups, the median duration of PPA was lowest among adolescents. The median duration of postpartum amenorrhea in 2016 was 1.1 months shorter among adolescents compared to young women.

**Figure 1.**
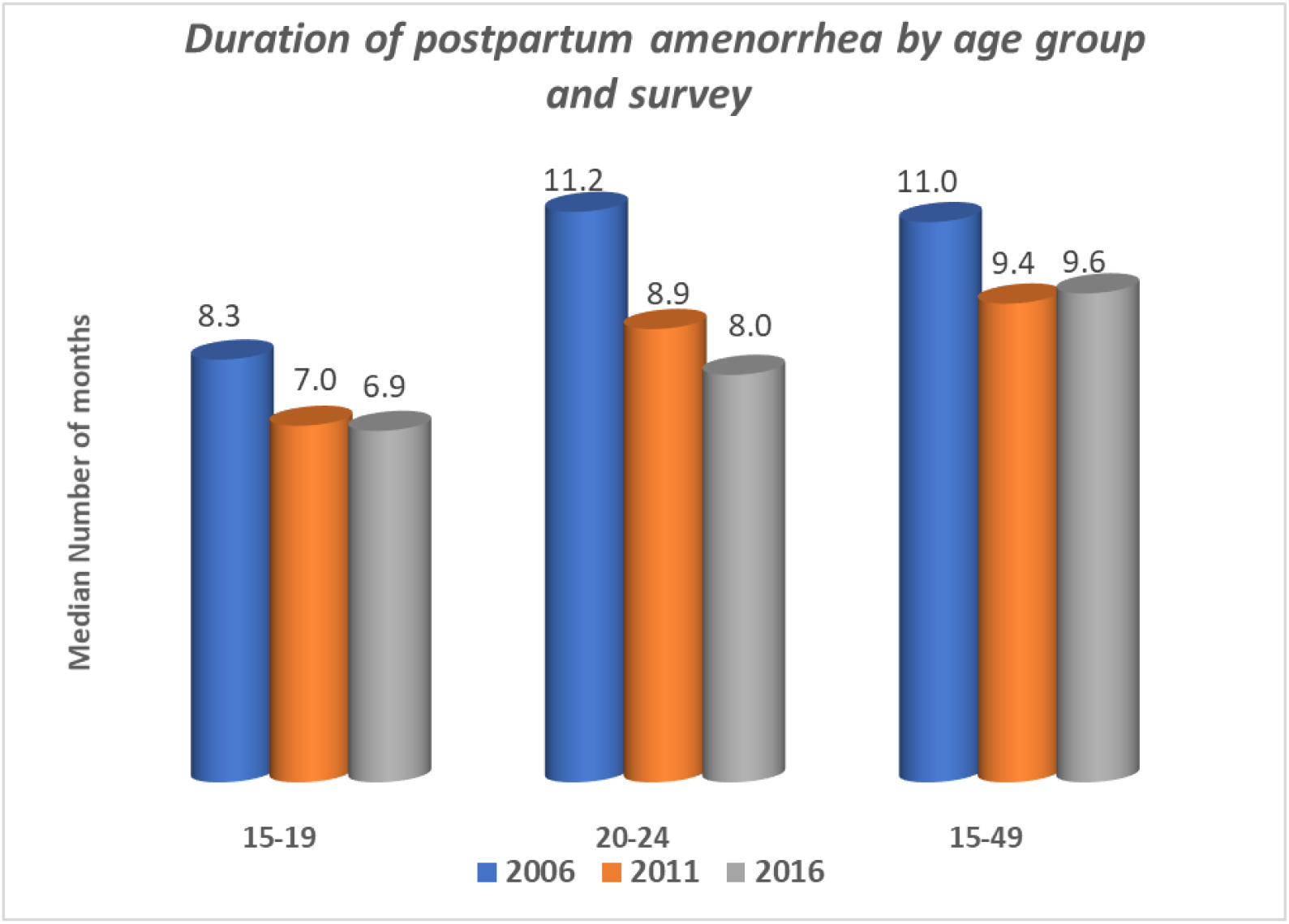
Duration of postpartum amenorrhea by age group and survey.

## DISCUSSION

This study explored the knowledge and use of lactational amenorrhea as a FP method among adolescents in Uganda. The findings indicate that nearly 60% of eligible adolescents in 2016 had knowledge about LAM, and there was an increase in the percentage of adolescents with knowledge of LAM in the examined period. The reported and correct use of LAM among eligible adolescents was very low, and there was a high but declining prevalence of passive LAM use. The results also show a decrease in the median duration of postpartum amenorrhea among adolescents between 2006 and 2016.

We found large increases in knowledge of LAM among all and eligible adolescents, especially between 2011 and 2016. Part of this increase may be attributed to changes in the measurement of LAM knowledge in 2016 where a description of LAM was read out to the respondents by survey enumerators, which was not the case in the years before. While some views have previously considered this as a quality concern resulting in misreporting by women, other analyses have supported the inclusion of the LAM description, as higher proportions of self-reported LAM users who correctly practised LAM were observed in surveys with the description (Fabic and Choi, 2013). Any true increase in levels of knowledge about LAM may be attributed to various efforts that have been implemented over time by the Ugandan government including free primary education, family planning initiatives including the use of community health workers, and promoting the provision of adolescent and youth friendly services (Atuyambe et al., 2015, Nalwadda et al., 2019). The percentage of eligible adolescents who were aware of LAM in this study was higher than what has been reported among women of reproductive age in other SSA countries (Abraha et al., 2018, Alvey, 2015, Ekpenyong et al., 2013). This may be explained by differences in the methods for determining knowledge of LAM. For example, studies by Abraha et al (2018) and Ekpenyong et al (2013) went beyond the DHS approach and included an assessment of respondent’s understanding of the three criteria for LAM practice. Regardless, the observed increase in knowledge may result in better attitudes towards and acceptability of LAM as an effective contraceptive method, which would contribute to increased adoption. But this would be dependent on the appropriateness of the information to which adolescents are exposed and their ability to apply it correctly, among other factors.

Findings from this study show very low percentages of eligible adolescents reporting the use of LAM as their primary FP method, and those that were correctly using it. The low prevalence of reported LAM use may partly be explained by methodological limitations posed by the DHS where if a woman reports use of both LAM and other modern methods, the most effective method is recorded and the woman is not recorded as a LAM user (Fabic and Choi, 2013). The use of other FP methods besides LAM during the first 6 months after childbirth has been reported (Afifi, 2007). This may explain the observed high proportion of women that were passively using LAM (meet the LAM criteria but do not report LAM use). The study findings relate to previous studies that also show a low prevalence of LAM use among women of reproductive age in general (Fabic and Choi, 2013) and adolescents in particular (Figaroa et al., 2020). Low correct use of LAM is a result of not fulfilling one or more of the three criteria, which also influence the effectiveness of LAM as a FP method. This would mean that, if new adolescent mothers are not correctly using LAM and they are not using any other FP methods within the first 6 months after childbirth, they would be at higher risk of becoming pregnant again. This would also vary based on factors like sexual activity or access to clinical or social support during this period. In other studies, violation of the exclusive breastfeeding and postpartum amenorrhea criteria have been reported to be the primary factors affecting the correct use of LAM (Van der Wijden and Manion, 2015, Sipsma et al., 2013). This is particularly important in adolescents among whom, early initiation and exclusive breastfeeding have been reported to be low, with a higher likelihood of discontinuation (Benova et al., 2020, Jara-Palacios et al., 2015).

Postpartum amenorrhea is one of the key determinants for correct LAM use and its effectiveness. The study findings showed a decrease in the median duration of PPA among adolescents from 8.3 (2006) to 6.9 (2016). The resumption of menses is a commonly used reported determinant of FP use among postpartum women (Gahungu et al., 2021), yet variably understood as reported by Cooper et al (2019), who found misconceptions about the return of fecundity in Tanzania (Cooper et al., 2019). The decreasing duration of PPA points to the need to strengthen postpartum FP among adolescents, as the risk of becoming pregnant may be higher in this sub-population. The review by Figaroa and colleagues noted wide variation in PPA duration among adolescents in LMICs (Figaroa et al., 2020); while a study in India among women with at least one livebirth reported a lower duration of PPA (5.7 months) than what was found in the present study (Singh et al., 2012). Breastfeeding practices have also been linked to a shorter total duration of PPA (Singh et al., 2012). According to the authors, the risk of resumption of menses decreases with the increasing duration of breastfeeding. This may explain the decreasing duration of PPA among adolescents and why it was lower compared to young women in this study.

## Limitations

This study benefitted from three rounds of highly comparable nationally representative data collected over ten years in Uganda to provide an understanding of patterns of LAM use among adolescents. We constructed several indicators of LAM knowledge and use, with sensitivity to the population under consideration (i.e. denominators). Nonetheless, this study has limitations. First, it was not possible to investigate factors associated with LAM knowledge or use among adolescents because of the small sample size. Such information would help inform strategies to improve the use of LAM in this key sub-population. The findings are also limited by the use of self-reported data on the use of LAM and its criteria especially exclusive breastfeeding and return of menses. Correct/accurate reporting on such variables is rather subjective and is affected by the individual’s understanding which is often limited. Also, the scope of knowledge of LAM assessed in the surveys is limited and may not provide a proper indication of the understanding of LAM among adolescents. Further qualitative studies can help elucidate the understanding and considerations for use of LAM in this important population segment.

## CONCLUSIONS

There is increasing awareness about LAM but, the reported and correct use of LAM is low among adolescents with a live baby under 6 months of age in Uganda. There is need to improve uptake of contraceptive methods especially in postpartum adolescents through targeted counselling and educational programs. This will go a long way in not only reducing their short-interpregancy intervals but improving their health and wellbeing including that of their children. Further, additional research is needed to better understand the dynamics of correct LAM use and transition to other FP methods among adolescent mothers.

## Data Availability

Data are available in a public, open access repository. Data are available for research purposes on www.dhsprogram.com after registration

## Acknowledgements

CB and PB would like to thank the Institute of Tropical Medicine in Belgium for the training that contributed to the development of this paper. We would also like to convey our thanks to the Demographic and Health Survey program for granting access to the survey datasets which were used in this study.

## Contributions

CB and PB share first authorship and contributed equally to this paper. CB, PB conceptualized the study, conducted data analysis and developed the manuscript. LB provided analysis guidance and supported the conceptualization process and data analysis. CB, PB and LB wrote the first manuscript draft. STW supported data analysis and participated in manuscript review. PW provided technical guidance to the manuscript development process. All authors reviewed the manuscript and approved the final manuscript.

## Patient consent for publication

Not required.

## Patient and public involvement

Patients or the public were not involved in the design, or conduct, or reporting, or dissemination plans of this research.

